# Subphenotypes of youth-onset type 2 diabetes mellitus and their association with distal symmetrical polyneuropathy

**DOI:** 10.1101/2024.10.01.24314707

**Authors:** Jiali Guo, Zhongyu Li, Rodrigo M. Carrillo-Larco, Daniel S. Hsia, Jessica L Harding, Mohammed K. Ali, Jithin Sam Varghese

## Abstract

**Context:** Individuals with youth-onset type 2 diabetes mellitus (T2DM) display substantial, but unexplained, heterogeneity in their clinical presentations and risk of complications such as diabetic neuropathy. Data-driven clustering may be useful in characterizing this heterogeneity.

**Objective:** To identify data-driven subphenotypes of newly diagnosed youth-onset T2DM and study their association with distal symmetric polyneuropathy (DSPN) at time of diagnosis.

**Design:** Cross-sectional

**Setting:** USA

**Participants:** 641 individuals with newly diagnosed T2DM aged 10-19 years from the SEARCH for Diabetes in Youth Study and the Treatment Options for Type 2 Diabetes in Adolescents and Youth (TODAY) study.

**Exposure(s):** Body mass index, HbA1c, fasting C-peptide, systolic blood pressure, diastolic blood pressure, LDL cholesterol and HDL cholesterol

**Main Outcome Measures:** Data-driven subphenotypes were identified from k-means clustering. The cross-sectional association of subphenotypes with DSPN, based on expert examination scores (≥2.5) from the Michigan Neuropathy Screening Instrument, were assessed using Poisson regressions with robust standard errors.

**Results:** Among 641 youth-onset T2DM, 58.2% were female, with 38.2% of participants ≤13 years having average BMI of 34.5 kg/m^2^ (SD: 6.5 kg/m^2^), and average HbA1c of 6.1% (IQR: 5.6-7.0). Three youth-onset subphenotypes were identified: mild obesity related diabetes (yMOD, 48.5%), severe insulin deficient diabetes (ySIDD, 18.7%) and severe insulin resistant diabetes (ySIRD, 32.7%). After adjusting for covariates, the prevalence of abnormal DSPN were 2.58 (95%CI: 1.74, 3.81) and 2.02 (95%CI: 1.40, 2.93) times among those classified as the ySIDD and ySIRD subphenotypes, relative to the yMOD subphenotype.

**Conclusions:** Youth-onset T2DM consisted of heterogeneous clinical subphenotypes with differences prevalence of DSPN. Management of youth-onset T2DM may need to consider strategies tailored to each subphenotype.

## Introduction

Youth-onset type 2 diabetes (T2DM) is rapidly increasing in prevalence in the United States with 12.5 cases per 100,000 individuals aged 10 to 19 years.^1,2^ The pathophysiology involves inadequate insulin secretion compensating for insulin resistance from puberty and excess adiposity, often among those with a family history of diabetes.^3,4,5^ Previous research suggests that youth-onset T2DM exists on a spectrum with heterogeneity in the rate of decline of beta-cell function, response to treatment, and risk of complications by sex, baseline glycemic control, race-ethnicity, and age at diagnosis.^4^ A precision medicine approach, which recognizes the observed heterogeneity of youth-onset T2DM, could therefore inform diagnosis and treatment interventions.^6^

The heterogeneity in clinical presentation and risk of complications of youth-onset T2DM are driven by a complex pathophysiology of glucotoxicity, insulin resistance, and lipotoxicity, that existing classifications do not address.^7,8^ First, patients exist on a spectrum of insulin resistance and secretion that not captured by current subgroups based on presence or absence of islet antibodies and abdominal obesity.^9,10^ Second, existing classifications are not useful for risk stratification since they do not consider differences in prognosis of youth-onset T2DM. For instance, youth-onset T2DM patients display a high burden of microvascular complications such as peripheral neuropathy with nearly 35% cumulative incidence at 15 years after diagnosis,^1,11^ which may depend on factors like age, BMI, inflammation, and medications (e.g., insulin). Moreover, characterizing the heterogeneity in clinical presentation of youth-onset T2DM can be improved by considering the extent of dysglycemia, high blood pressure and high cholesterol at diagnosis, which are associated with elevated risk of complications.

Recent studies of data-driven classification of newly diagnosed cases of diabetes (T1DM, latent autoimmune diabetes, and T2DM) have identified five frequently replicated subgroups (i.e., subphenotypes): mild age-related diabetes (MARD), mild obesity-related diabetes (MOD), severe insulin resistant diabetes (SIRD), severe insulin-deficient diabetes (SIDD), and severe autoimmune diabetes (SAID), based on six variables (GAD antibodies, HbA1c, BMI, age, HOMA2-B, HOMA2-IR).^12,13^ These subphenotypes are associated with differences in response to treatments and risk of distal symmetric polyneuropathy (DSPN).^14^ For example, relative to other subphenotypes, SIDD and SIRD were associated with the highest risk of DSPN. Therefore, using data of the two largest studies of youth-onset T2DM from the USA, our objectives were to 1) identify data-driven subphenotypes of newly diagnosed youth-onset T2DM; and 2) to study their associations with DSPN at T2DM diagnosis. Findings from this study can improve risk stratification of newly diagnosed youth-onset T2DM cases.

## Materials and Methods

### Study Population

#### SEARCH

The SEARCH for Diabetes in the Youth Study consists of a registry of existing and newly diagnosed diabetes (T1DM, T2DM, other types, or hybrids) who were 19 years or younger, from six clinical centers in Ohio, Colorado, Washington, South Carolina, Southern California and Hawaii.^10^ The first four sites identified cases from a geographically defined population, while the latter two identified participants from the membership of a participating health plan, i.e. Kaiser Permanente. SEARCH consists of four phases of enrollment (Phase 1: 2000-05, Phase 2: 2005-10, Phase 3: 2010-15, Phase 4: 2015-20). Cross-sectional data from enrollment was used for each participant in this study.

#### TODAY

The Treatment Options for Type 2 Diabetes in Adolescents and Youth (TODAY) study was a multicenter, randomized control trial of metformin monotherapy, compared to alternative approaches (metformin + rosiglitazone, metformin + lifestyle intervention) among newly diagnosed T2DM (<2 years and autoantibody negative) for individuals aged 10-17 years who did not require insulin and were overweight or obese (BMI > 85^th^ percentile).^15^ Duration of diabetes was ≤5 months for 48% of participants. Cross-sectional data from the enrollment wave before intervention allocation were used for this analysis.

#### Type 2 Diabetes Diagnostic Criteria

Diabetes cases in the SEARCH study were validated through one of two primary criteria: a physician’s diagnosis of diabetes, or self-reporting of a physician’s diagnosis by the patient (or their parent) during an interview or survey. A physician diagnosis was considered confirmed if it met any of the following conditions: medical record review that reveals a physician’s diagnosis of diabetes; direct verification of the diagnosis, or a referral to the study by a clinician; listing of diabetes as an underlying or contributing cause of death on a death certificate; inclusion in a clinical database that mandates clinician verification of the diagnosis.

T2DM cases in the TODAY study were diagnosed according to the American Diabetes Association (ADA) criteria,^16^ with confirmation via documented medical records. The diagnosis was established based on one or more laboratory parameters: fasting glucose ≥126 mg/dL, random glucose ≥200 mg/dL, or a two-hour oral glucose tolerance test (OGTT) glucose ≥200 mg/dL. For patients with normal fasting glucose and elevated two-hour glucose levels during the OGTT, the HbA1c must be ≥6% as per the study protocol.^17^ Additionally, for patients who had been previously diagnosed with diabetes and were on medication at the time of screening for eligibility in this study, HbA1c ≥8% at the time of their initial diagnosis was considered sufficient evidence of eligibility if no documented laboratory determination of serum glucose was available.

#### Analytic Sample

We included all newly diagnosed participants aged 10 to 19 years. Newly diagnosed diabetes was defined as a duration of diabetes of ≤5 months at enrollment in TODAY, and ≤1 year in SEARCH. Participants from the SEARCH study were included if they were not classified as type 1 diabetes or Maturity Onset Diabetes of the Young (MODY), i.e. they were classified as type 2 or unknown form of diabetes based on etiologic evidence by expert adjudicators (**Supplementary Figure 1**). We excluded participants who were missing body mass index (BMI) at enrollment in either study. Additionally, we excluded participants for whom all blood-based laboratory biomarker assessments were unavailable. Our final analytic sample consisted of 641 participants with newly diagnosed youth-onset T2DM from both TODAY (n=337) and SEARCH (n=304) studies.

### Data collection and variable specification

#### Clinical characteristics of youth-onset T2DM

We extracted relevant socio-demographics and clinical characteristics of the analytic sample for both studies. Socio-demographic data included sex, age (harmonized across studies as ≤13, 14-15, >15 years based on data sharing policies), and race and ethnicity (Non-Hispanic Black, Non-Hispanic White, Hispanic, Non-Hispanic Other). Clinical characteristics included vitals such as BMI, systolic and diastolic blood pressure; laboratory parameters such as hemoglobin A1c (HbA1c), fasting C-peptide, fasting glucose, fasting insulin, total cholesterol, low-density lipoprotein (LDL) cholesterol, high-density lipoprotein (HDL) cholesterol, and triglyceride levels (measurement protocols in **Supplementary Table 1**); and self-reported usage of insulin among SEARCH participants. Since the data of TODAY study were extracted from the enrollment wave, we did not use the information on treatment group assignment.

#### Distal symmetrical polyneuropathy

Cross-sectional assessment of DSPN was conducted using the validated Michigan Neuropathy Screening Instrument (MNSI) for both SEARCH and TODAY.^18^ The MNSI measurement consisted of two separate assessments, an expert examination and a self-administered questionnaire. During the 4-item expert examination that included inspection and assessment of vibratory sensation and ankle reflexes, a health professional inspects each foot for deformities, dry skin, calluses, infections and fissures. The total possible score of all examinations is 8 points and, in the published scoring algorithm, a score ≥2.5 is considered abnormal and indicative of DSPN.^19^ Additionally, the self-administered questionnaire consists of 15 questions, where each response counts as one point. Responses to impaired circulation and general asthenia were excluded in the published scoring algorithm (≥4 indicative of DSPN).^19^

### Statistical Analysis

#### Data-driven clustering to identify subphenotypes of T2DM

We calculated age-adjusted variables for clustering as residuals from the linear regression of each of these seven variables on age.^20^ We used these variables to perform an exploratory k-means cluster analysis (k: 2 to 10). We identified the optimal number of clusters using a combination of an elbow plot (**Supplementary Figure 2**) and clinically meaningful sample sizes within each cluster across different numbers of clusters. This approach determined that three clusters represented the optimal solution for classifying the analytic sample into subphenotypes. We compared clinical characteristics between the clusters to inform our labeling of subphenotypes. To compare our findings with existing classifications based on adiposity, we compared the overlap of the three subphenotypes with tertiles of BMI.^10^ To account for missingness (<5%) in variables that were used to identify the data-driven clusters (i.e., BMI, HbA1c, fasting C-peptide, systolic blood pressure, diastolic blood pressure, LDL cholesterol, or HDL cholesterol) and missing indicators in the MNSI examination, we used a non-parametric single imputation method (k-nearest neighbors; k = 5). Clusters were then labelled after comparing their clinical characteristics and their similarities to adult-onset subphenotypes.

#### Association of subphenotypes with DSPN

We conducted a cross-sectional analysis of the pooled dataset of SEARCH and TODAY. We excluded individuals who did not have any MNSI measurement at the time of inclusion in the study and accounted for missing DSPN indicators using inverse probability weights.^21^ We estimated the association of membership in subphenotypes with DSPN (as defined by abnormal examination scores), and used yMOD as the reference group, adjusting for age, sex and race-ethnicity. We used modified Poisson regressions to estimate prevalence ratios and 95% robust confidence intervals.

#### Sensitivity Analysis

First, to study if the identification protocol for youth-onset T2DM in SEARCH may have biased our results, we repeated the analysis using two alternative definitions provided by the SEARCH investigators: a) cases with negative or missing diabetes-associated autoantibodies, and b) an initial provider classification including all known T2DM and cases of unknown etiology. Next, to address the potential bias from missing MNSI data, we excluded cases lacking DSPN indicators and conducted a sensitivity analysis focusing exclusively on complete cases. Finally, given that the TODAY study used highly restrictive inclusion criteria of non-insulin dependent T2DM, and considering the broader diversity within the SEARCH study, we repeated the analysis using only the SEARCH dataset.

All analysis was carried out using R 4.3.3 and Python 3.12.4.

## Results

Our analytic sample consisted of 641 participants aged 10-19 years with newly diagnosed T2DM. Of these 304 (47.4%) were from the SEARCH cohort study, and 337 (52.6%) were from the TODAY trial. The study population were 58.2% female with 38.2% of participants ≤13 years of age at T2DM diagnosis. Non-Hispanic White, non-Hispanic Black, and Hispanic individuals each accounted for approximately 30% of the sample, and non-Hispanic other races made up 8.7% of the sample. Individuals excluded from the analytic sample (n = 77) for missing DSPN indicators in SEARCH were more likely to be ≤13 years (67.5% versus 35.5%) and Non-Hispanic White (64.9% versus 32.2%) (**Supplementary Table 2**).

The average BMI of the study population was 34.5 kg/m^2^ (SD: 6.5 kg/m^2^). The mean HbA1c level was 6.1% (IQR: 5.6%, 7.0%), while the average systolic and diastolic blood pressures were 114.2 mmHg (SD: 11.6 mmHg) and 68.8 mmHg (SD: 9.7 mmHg), respectively (**Table 1**). Descriptive characteristics stratified by sex for SEARCH and TODAY are provided in **Supplementary Table 3**. Missing data for variables used in clustering were <5% (**Supplementary Table 4**).

**Table 1.** Descriptive characteristics of the analytic sample stratified by identified subphenotypes.

|  | **Total** | **yMOD** | **ySIDD** | **ySIRD** |
| --- | --- | --- | --- | --- |
| *N* | 641 | 311 (48.5%) | 120 (18.7%) | 210 (32.7%) |
| SEARCH | 304 (47.4%) | 86 (27.7%) | 94 (30.9%) | 124 (40.8%) |
| TODAY | 337 (52.6%) | 225 (72.3%) | 26 (7.7%) | 86 (25.5%) |
| Age at diagnosis (years) |  |  |  |  |
| $\leq 13$ | 245 (38.2%) | 120 (38.6%) | 48 (40%) | 77 (36.7%) |
| 14-15 | 146 (22.8%) | 72 (23.2%) | 28 (23.3%) | 46 (21.9%) |
| $> 15$ | 250 (39%) | 119 (38.3%) | 44 (36.7%) | 87 (41.4%) |
| Female % | 641 (58.2%) | 311 (60.8%) | 120 (60%) | 210 (53.3%) |
| Race-Ethnicity |  |  |  |  |
| *NH White* | 172 (26.8%) | 77 (24.8%) | 41 (34.2%) | 54 (25.7%) |
| *NH Black* | 205 (32%) | 80 (25.7%) | 42 (35%) | 83 (39.5%) |
| *Hispanic* | 208 (32.4%) | 127 (40.8%) | 27 (22.5%) | 54 (25.7%) |
| *NH Other* | 56 (8.7%) | 27 (8.7%) | 10 (8.3%) | 19 (9%) |
| ***Key biomarkers for clustering*** |  |  |  |  |
| BMI (kg/m<sup>2</sup>) | 34.5 (6.5) | 33.5 (3.7) | 29.3 (6.1) | 38.8 (7.3) |
| HbA1c (%) | 6.1 (5.6, 7) | 5.7 (5.4, 6.2) | 7.6 (6.3, 8.9) | 6.2 (5.8, 7.4) |
| Fasting C-peptide (ng/mL) | 3.8 (1.8) | 3.5 (1.4) | 2.1 (1.1) | 5.1 (1.8) |
| LDL cholesterol (mg/dL) | 90.7 (27.9) | 80.2 (22.1) | 109.4 (30.1) | 95.7 (27.5) |
| HDL cholesterol (mg/dL) | 39.2 (9.2) | 37.1 (7.3) | 48.2 (9.6) | 37.2 (8.3) |
| Systolic BP (mmHg) | 114.2 (11.6) | 109.3 (9) | 109.7 (10.2) | 124 (9.4) |
| Diastolic BP (mmHg) | 68.8 (9.7) | 64.2 (7.4) | 67.8 (8.2) | 76.2 (9.1) |
| ***Other biomarkers*** |  |  |  |  |
| Fasting glucose (mg/dL) <sup>a</sup> | 106.5 (21.3) | 102 (18.2) | 117.3 (25.9) | 114.8 (24) |
| Fasting Insulin (uU/mL) <sup>a</sup> | 30.1 (22.2) | 26.2 (18.4) | 20 (10.3) | 43.4 (27.8) |
| Total cholesterol (mg/dL) | 154.8 (36.1) | 140.2 (27.9) | 181.7 (36.6) | 162.3 (36.3) |
| Triglycerides (mg/dL) <sup>a</sup> | 107.7 (67.1) | 101 (57.5) | 82 (40.7) | 133.1 (87.1) |
| ***DSPN*** |  |  |  |  |
| MNSI Measured | 501 (78.2%) | 268 (53.5%) | 80 (16.0%) | 153 (30.5%) |
| Abnormal Examination | 113 (22.6%) | 32 (28.3%) | 32 (28.3%) | 49 (43.4%) |
| Abnormal Questionnaire | 67 (13.4%) | 29 (43.3%) | 8 (11.9%) | 20 (29.9%) |
| Abnormal Examination<br>or Abnormal<br>Questionnaire | 146 (29.1%) | 52 (35.6%) | 34 (23.3%) | 60 (41.1%) |

<sup>a</sup> Only available for TODAY (N = 337); yMOD: Youth-onset Mild Obesity-related Diabetes, ySIDD: Youth-onset Severe Insulin Deficient Diabetes, ySIRD: Youth-onset Severe Insulin Resistant Diabetes
Data are mean (SD) or median (IQR) for continuous variables and frequency (percentage) for categorical variables. Abnormal examination and abnormal questionnaire were defined as scores $\geq 2.5$ and 4 respectively.

### Identification of Subphenotypes

Three clusters were identified after k-means clustering of age-adjusted variables (**Supplementary Table 5**). Cluster 1, comprising 120 (18.7%) individuals, had lower BMI and fasting C-peptide but higher values of HDL cholesterol and HbA1c compared to the other subphenotypes, and was therefore labeled as youth-onset severe insulin deficient diabetes (ySIDD). Cluster 2, which included 311 (48.5%) out of the 641 participants, was characterized by higher BMI and lower HbA1c relative to ySIDD and was labeled as youth-onset mild obesity related diabetes (yMOD). Cluster 3, labeled as severe insulin resistant diabetes (ySIRD), containing 210 (32.7%) participants with highest values of BMI, fasting C-peptide, blood pressure, and LDL cholesterol, and lower values of HbA1c and HDL cholesterol compared to ySIDD (**Figure, Table 1**). Of all ySIDD cases, 72.5% belonged to the lowest BMI tertile while 63.3% of ySIRD cases belonged to the highest BMI tertile (**Supplementary Table 6**). Vitals and laboratory parameters of subphenotypes were similar when clustering was conducted separately by sex (**Supplementary Table 7**).

**Figure.**
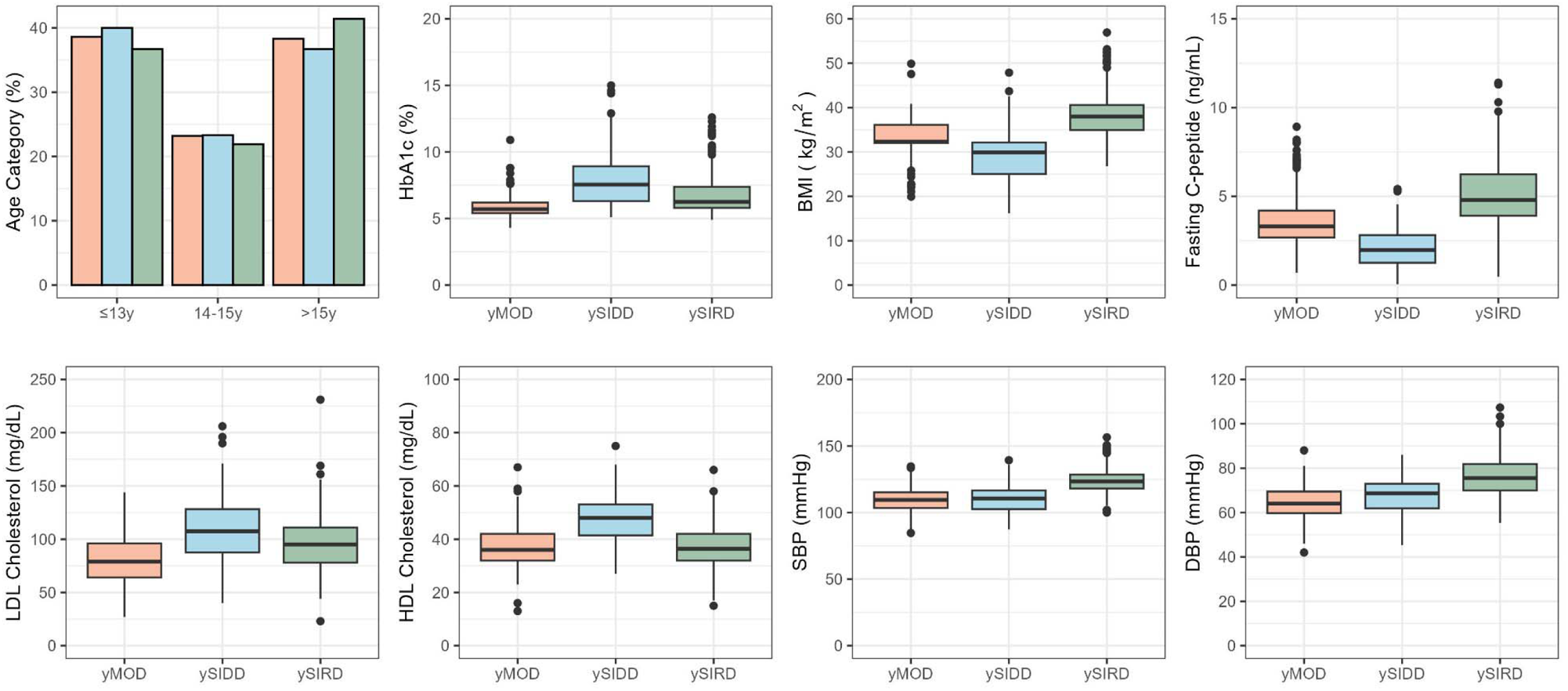
Distribution of clinical characteristics by subphenotype

### Association between subphenotypes and DSPN

Of the total analytic sample, 140 participants did not have any MNSI scores, and 14 participants had at least one missing item from the MNSI measurements at study inclusion (**Supplementary Table 8**). Relative to the yMOD subphenotype, the prevalence of individuals with DSPN based on examination scores were 2.73 (PR; 95%CI: 1.84, 4.04) and 2.30 (95%CI: 1.59, 3.34) times among those classified as the ySIDD and ySIRD subphenotypes respectively after accounting for non-response to MNSI examination (**Table 2**). After adjusting for age, sex and race/ethnicity, the prevalence of participants with abnormal DSPN based on the examination were 2.58 (95%CI: 1.74, 3.81) and 2.02 (95%CI: 1.40, 2.93) times among those classified as the ySIDD and ySIRD subphenotypes, relative to the yMOD subphenotype.

**Table 2.** Prevalence ratio of distal symmetrical polyneuropathy by subphenotype.

| **Prevalence ratio (95% CI)** | **Unadjusted** | **Adjusted** |
| --- | --- | --- |
| yMOD (reference) | Ref (1.00) | Ref (1.00) |
| ySIDD | 2.73 (1.84, 4.04) | 2.58 (1.74, 3.81) |
| ySIRD | 2.3 (1.59, 3.34) | 2.02 (1.4, 2.93) |

<sup>a</sup> Only available for SEARCH (N = 304);
yMOD: Youth-onset Mild Obesity-related Diabetes, ySIDD: Youth-onset Severe Insulin Deficient Diabetes, ySIRD: Youth-onset Severe Insulin Resistant Diabetes
Associations are prevalence ratios from marginal structural Poisson regressions using multiple imputation for missing covariates and inverse probability weights for non-participation in Michigan Neuropathy Screening Instrument assessment. Abnormal neuropathy score is adjusted for age category, female sex and race-ethnicity.

### Sensitivity Analysis

The sample based on cases with negative or missing autoantibodies consisted of 722 newly diagnosed T2DM cases aged 10–19 years. Among these, 46.8% belonged to the yMOD subphenotype, 22.2% were in the ySIDD subphenotype, and 31.0% were classified as the ySIRD subphenotype (**Supplementary Table 9**). Based on the initial provider diagnosis criterion, the analytic sample included 640 participants, with 46.7% classified as yMOD, 21.9% as ySIDD, and 31.4% as ySIRD (**Supplementary Table 10**). The means and SDs of the seven clustering variables were similar across different diagnostic criteria. We also did not observe any significant differences in the means and SDs of the clustering variables between our study sample (main clusters), the complete cases, and the clusters derived from clustering the SEARCH sample (**Supplementary Figure 4**). Prevalence of DSPN (relative to yMOD) were higher for ySIDD and ySIRD classified according to the autoantibody classification and initial provider classification (**Supplementary Table 11; Supplementary Figure 5**). However, for the SEARCH-only sample, the prevalence of DSPN among those classified as the ySIDD and ySIRD subphenotypes was similar to yMOD (**Supplementary Table 11**).

## Discussion

In this study, we identified three subphenotypes of youth-onset T2D based on laboratory parameters and vitals – yMOD, ySIDD and ySIRD. The latter two subphenotypes—ySIDD and ySIRD—showed a higher prevalence of abnormal DSPN symptoms at T2DM diagnosis compared to the yMOD subphenotype. This suggests that identifying these subphenotypes early could enhance risk stratification, as ySIDD and ySIRD patients appear to be more susceptible to DSPN. Therefore, implementing diagnostic algorithms in electronic health records to detect these subphenotypes and monitor DSPN risk can potentially lead to improved patient outcomes.

Consistent with our findings, most studies among adult-onset T2DM reported a higher proportion of individuals as the MOD subphenotype and lower proportion of adults classified as SIDD and SIRD subphenotypes.^22^ Despite differences in age of onset of 40 years or more in the adult studies, the clinical characteristics of adult-onset subphenotypes resembled those of youth-onset subphenotypes. For instance in the German Diabetes Study (GDS) and All New Diabetics in Scania (ANDIS) cohort from Sweden, BMI was higher among adult-onset SIRD and MOD, as compared with adult-onset SIDD (**Supplementary Table 12**).^12,14^ Furthermore, consistent with findings from these studies, HbA1c was higher among adult-onset SIDD, relative to adult-onset SIRD and adult-onset MOD. Additionally, we observed greater burden of DSPN among the ySIDD and ySIRD subphenotypes, relative to yMOD, consistent with evidence from adult-onset SIDD and SIRD in the GDS.^14^

The increasing incidence of DSPN among children and youth with type 1 diabetes was previously highlighted, and is associated with higher risk of ulcers and non-traumatic amputation.^23^ Our findings suggest add to the evidence that burden of DSPN among youth-onest T2DM may be further exacerbated by obesity, ^24^ based on membership in ySIDD and ySIRD subphenotypes. Furthermore, reports from TODAY and SEARCH studies suggest that rates of DSPN in youth-onset T2DM are higher than those in T1DM, comparable to adult-onset T2DM.^25–27^ However, nearly 50% of DSPN are also asymptomatic with contributing factors such male sex, poor glycemic control, duration of diabetes, high BMI, low HDL cholesterol, and smoking.^25,26^ Despite this, the underlying etiology and strategies for preventing or managing DSPN in children and youth with diabetes remain unclear. The variations in prevalence across the subphenotypes identified in this study can improve diagnosis and management through risk stratification.

Our study enhances the risk stratification of youth-onset T2DM using seven biomarkers to better capture the heterogeneity in clinical presentation.^10^ However, there are several limitations to consider. These findings should be interpreted as exploratory given the small sample size (N = 641) which may have left us underpowered to detect small and clinically meaningful differences between subphenotypes of T2DM, including in responses to treatment by intervention group in the TODAY study. Moreover, we were unable to incorporate the nuances of different study designs and mechanisms of participant selection of the SEARCH and TODAY studies in the clustering. For instance, the SEARCH cohort was not limited to T2DM, whereas the TODAY trial had more restrictive recruitment criteria, including non-insulin dependent T2DM, and we were unable to identify individuals with duration of youth-onset T2DM between 6 and 12 months. Nevertheless, our study provides preliminary data from the most comprehensive databases of youth-onset T2DM and documents the heterogeneity of T2DM.

### Conclusion

In summary, our study introduced a novel data-driven classification for youth-onset T2DM, underscoring a higher burden of DSPN among some subphenotypes that may be useful in defining a precision medicine approach to managing youth-onset T2DM to prevent morbidity from complications. Further research is needed to validate these findings in larger and more diverse longitudinal datasets. Additionally, longitudinal studies are essential for tracking the development of DSPN in individuals who are initially free at diagnosis of youth-onset T2DM and for exploring the potential advantages of tailored treatment strategies for these novel subphenotypes.

## Abbreviations

MNSI: Michigan Neuropathy Screening Instrument
T1DM: Type 1 Diabetes
T2DM: Type 2 Diabetes
DAA: Diabetes Associated Antibody
DSPN: Distal Symmetric Polyneuropathy
yMOD: Youth-onset Mild Obesity-related Diabetes
ySIDD: Youth-onset Severe Insulin-Deficient Diabetes
ySIRD: Youth-onset Severe Insulin-Resistant Diabetes

## Ethics approval and consent to participate

We were exempt from ethical approval by the Institutional Review Board of Emory University. All participants gave written informed consent before participation in both SEARCH and TODAY studies.

## Data availability statement

The code for the analysis is available on https://github.com/jvargh7/diabetes_subphenotypes_youth. Data for SEARCH and TODAY are available from the NIDDK Biorepository.

## Consent for publication

Not applicable

## Author contributions

JSV, ZL and JG conceptualized the study. JG conducted the analysis and wrote the first draft with inputs from ZL and JSV. All authors reviewed and edited subsequent drafts.

