## Supplementary Materials for "Subphenotypes of youth-onset type 2 diabetes mellitus and their association with distal symmetrical polyneuropathy"

**Supplementary Table 1. Comparison of analytical methods for laboratory and anthropometry parameters**

|  | **SEARCH** | **TODAY** |
| --- | --- | --- |
| **Height** | Height and weight are measured during a physical examination conducted by trained medical staff at the baseline in-person visit. The examination also includes waist measurements, heart rate, and blood pressure. | Height and weight are measured at each study visit for participants. Height and weight are also collected from the family support person (FSP) annually to assess the impact of the FSP's weight loss or participation in the lifestyle intervention on the participant's anthropometrics and body composition. |
| **Weight** |  |  |
| **HbA1c** | HbA1c is measured by taking a blood sample during an early morning appointment, after the participant has fasted for 10 hours. This measure provides an assessment of long-term blood sugar control. | HbA1c is measured by performing baseline and interval assays at a central laboratory. It serves as the primary outcome for defining treatment failure, with values ≥ 8% over a 6-month period indicating failure. Investigators and participants are blinded to HbA1c results, except when alerts are provided if HbA1c exceeds certain thresholds to ensure adherence and safety. |
| **LDL Cholesterol** | LDL cholesterol levels are measured from the blood samples collected during the baseline in-person visit. These blood samples are used to assess various health markers, including different types of cholesterol, to monitor the participant's metabolic status. | LDL cholesterol is measured as part of the screening process to determine eligibility, along with triglycerides (TG). Additional lipid assays, including free fatty acids, lipoprotein subclass levels, average LDL particle density, and total ApoB levels, are measured from blood samples collected at baseline, six months, annually, the primary endpoint, and the end of the study. |
| **Fasting C-peptide** | Fasting C-peptide is measured under conditions of metabolic stability, defined as no episodes of diabetic ketoacidosis (DKA) in the month prior to the test. Participants provide a blood sample after fasting overnight, which is then analyzed to determine the concentration of C-peptide. This measurement helps assess insulin production and classify the type of diabetes | Fasting C-peptide is measured by collecting fasting blood samples from participants. These samples are then analyzed to determine C-peptide levels, which are used to assess insulin secretion. This measurement is taken at baseline, 6 months, annually, at the primary endpoint, and at the end of the study. |
| **Systolic Blood Pressure** | Blood pressure (SBP and DBP) is measured at every visit using a standard protocol. Measurements are taken while the participant is seated, with the arm supported at heart level, after a rest period of 5 minutes. The appropriate cuff size is used to ensure accurate readings, and multiple readings are taken to calculate an average value for SBP and DBP. | Blood pressure (SBP and DBP) is measured at every study visit using standard clinical procedures. Measurements are taken at baseline, every two months during the first year, and then quarterly thereafter. These repeated measurements help monitor changes in blood pressure throughout the study duration and ensure consistency across different clinical centers. |
| **Diastolic Blood Pressure** |  |  |

**Supplementary Table 2. Comparison of analytic and excluded sample from complete-case analysis**

|  | **SEARCH** |  | **TODAY** |  |
| --- | --- | --- | --- | --- |
|  | **Included** | **Excluded** | **Included** | **Excluded** |
| *N* | 304 (47.4%) | 77 (12.0%) | 337 (52.6%) | 0 |
| Socio-demographics |  |  |  |  |
| Age at diagnosis |  |  |  |  |
| *<=13* | 108 (35.5%) | 52 (67.5%) | 137 (40.7%) | - |
| *14-15* | 43 (14.1%) | 8 (10.4%) | 103 (30.6%) | - |
| *>15* | 153 (50.3%) | 17 (22.1%) | 97 (28.8%) | - |
| Female | 304 (55.9%) | 77 (61%) | 337 (60.2%) | - |
| Race-Ethnicity |  |  |  |  |
| *NH White* | 98 (32.2%) | 50 (64.9%) | 74 (22%) | - |
| *NH Black* | 112 (36.8%) | 12 (15.6%) | 93 (27.6%) | - |
| *Hispanic* | 67 (22%) | 12 (15.6%) | 141 (41.8%) | - |
| *NH Other* | 27 (8.9%) | 3 (3.9%) | 29 (8.6%) | - |
| BMI (kg/m^2^) | 34.4 (9.1) | 22.4 (5) | 34.5 (2.6) | - |
| HbA1c (%) | 6.6 (5.8, 8.3) | - | 5.9 (5.4, 6.3) | - |
| Insulin Usage | 382 (59.2%) | 76 (89.5%) | - | - |
| Fasting glucose (mg/dL) ^a^ | - | - | 106.5 (21.3) | - |
| Fasting Insulin (uU/mL) ^a^ | - | - | 30.1 (22.2) | - |
| Fasting C-peptide (ng/mL) | 3.8 (2.2) | - | 3.8 (1.6) | - |
| LDL cholesterol (mg/dL) | 99.1 (30.7) | - | 83.4 (23.9) | - |
| HDL cholesterol (mg/dL) | 39.5 (9.5) | - | 38.9 (9) | - |
| Total cholesterol (mg/dL) | 168.3 (39.3) | - | 143.4 (28.6) | - |
| Triglycerides (mg/dL) ^a^ | - | - | 107.7 (67.1) | - |
| Systolic BP (mmHg) | 115.5 (12.5) | 107 (11.8) | 113 (10.7) | - |
| Diastolic BP (mmHg) | 71.2 (10.4) | 67.3 (8.8) | 66.7 (8.6) | - |

^a^ Only available for TODAY (N = 337); yMOD: Youth-onset Mild Obesity-related Diabetes, ySIDD: Youth-onset Severe Insulin Deficient Diabetes, ySIRD: Youth-onset Severe Insulin Resistant Diabetes

**Supplementary Table 3. Descriptive characteristics by sex**

|  | **SEARCH** |  | **TODAY** |  |
| --- | --- | --- | --- | --- |
|  | **Male** | **Female** | **Male** | **Female** |
| *N* | 134 (20.9%) | 170 (28.4%) | 134 (26.5%) | 203 (28.1%) |
| Cluster |  |  |  |  |
| *yMOD* | 34 (25.4%) | 52 (30.6%) | 88 (65.7%) | 137 (67.5%) |
| *ySIDD* | 41 (30.6%) | 53 (31.2%) | 7 (5.2%) | 19 (9.4%) |
| *ySIRD* | 59 (44%) | 65 (38.2%) | 39 (29.1%) | 47 (23.2%) |
| Socio-demographics |  |  |  |  |
| Age at diagnosis |  |  |  |  |
| *<=13* | 36 (26.9%) | 72 (42.4%) | 38 (28.4%) | 99 (48.8%) |
| *14-15* | 19 (14.2%) | 24 (14.1%) | 49 (36.6%) | 54 (26.6%) |
| *>15* | 79 (59%) | 74 (43.5%) | 47 (35.1%) | 50 (24.6%) |
| Race-Ethnicity |  |  |  |  |
| *NH White* | 53 (39.6%) | 45 (26.5%) | 35 (26.1%) | 39 (19.2%) |
| *NH Black* | 36 (26.9%) | 76 (44.7%) | 34 (25.4%) | 59 (29.1%) |
| *Hispanic* | 31 (23.1%) | 36 (21.2%) | 56 (41.8%) | 85 (41.9%) |
| *NH Other* | 14 (10.4%) | 13 (7.6%) | 9 (6.7%) | 20 (9.9%) |
| BMI (kg/m^2^) | 33.4 (9.8) | 35.1 (8.5) | 34.6 (2.6) | 34.5 (2.6) |
| HbA1c (%) | 6.8 (6, 8.4) | 6.4 (5.8, 8.1) | 5.8 (5.4, 6.2) | 5.9 (5.5, 6.4) |
| Insulin Usage | 132 (53.8%) | 170 (51.2%) | - | - |
| Fasting glucose (mg/dL) ^a^ | - | - | 107.9 (22.4) | 105.5 (20.6) |
| Fasting Insulin (uU/mL) ^a^ | - | - | 30.3 (26.3) | 30 (19.1) |
| Fasting C-peptide (ng/mL) | 3.5 (2.3) | 3.9 (2) | 3.7 (1.5) | 3.9 (1.6) |
| LDL cholesterol (mg/dL) | 98.1 (29.3) | 99.5 (30.3) | 83.6 (24.7) | 83.3 (23.4) |
| HDL cholesterol (mg/dL) | 38.2 (10.3) | 40.7 (8.4) | 36.8 (8.2) | 40.3 (9.3) |
| Total cholesterol (mg/dL) | 168 (38.4) | 168.5 (40.1) | 141.3 (29.7) | 144.8 (27.9) |
| Triglycerides (mg/dL) ^a^ | - | - | 108.5 (74.6) | 107.2 (61.7) |
| Systolic BP (mmHg) | 118 (12.3) | 113.5 (12.1) | 115.3 (10.5) | 111.5 (10.5) |
| Diastolic BP (mmHg) | 72.2 (10.5) | 70.3 (10.1) | 66.5 (8.2) | 66.8 (8.9) |

^a^ Only available for TODAY (N = 337); yMOD: Youth-onset Mild Obesity-related Diabetes, ySIDD: Youth-onset Severe Insulin Deficient Diabetes, ySIRD: Youth-onset Severe Insulin Resistant Diabetes

**Supplementary Table 4. Missingness of cluster variables by study, N = 641**

|  | Total | SEARCH | TODAY |
| --- | --- | --- | --- |
| BMI | 0 | 0 | 0 |
| HbA1c | 4 (0.6%) | 4 (1.3%) | 0 |
| Fasting C-peptide | 17 (2.7%) | 16 (5.3%) | 1 (0.3%) |
| SBP | 4 (0.6%) | 4 (1.3%) | 0 |
| DBP | 5 (0.8%) | 5 (1.6%) | 0 |
| LDL Cholesterol | 19 (3.0%) | 18 (5.9%) | 1 (0.3%) |
| HDL Cholesterol | 19 (3.0%) | 18 (5.9%) | 1 (0.3%) |

**Supplementary Table 5. Coefficients of age category for cluster variables**

| Age at diagnosis (years) | BMI | HbA1c | Fasting C-peptide | SBP | DBP | LDL Cholesterol | HDL Cholesterol |
| --- | --- | --- | --- | --- | --- | --- | --- |
| 14-15 | 1.53 (0.21, 2.85) | -0.01 (-0.33, 0.32) | 0.05 (-0.33, 0.43) | 5.28 (2.99, 7.57) | 2.53 (0.6, 4.47) | 0.48 (-5.21, 6.18) | -0.99 (-2.86, 0.89) |
| >15 | 2.29 (1.16, 3.43) | 0.4 (0.13, 0.68) | 0.17 (-0.16, 0.49) | 6.47 (4.5, 8.45) | 5.2 (3.54, 6.87) | 6.89 (1.99, 11.78) | -1.67 (-3.29, -0.06) |

**Supplementary Table 6. Distribution of BMI classification within subphenotypes**

|  | yMOD | ySIDD | ySIRD |
| --- | --- | --- | --- |
| Low BMI | 106 (34.1%) | 87 (72.5%) | 21 (10.0%) |
| Moderate BMI | 135 (43.4%) | 22 (18.3%) | 56 (26.7%) |
| High BMI | 70 (22.5%) | 11 (9.2%) | 133 (63.3%) |
| Total | 311 (48.5%) | 120 (18.7%) | 210 (32.7%) |

We classified BMI residuals into tertiles.

yMOD: Youth-onset Mild Obesity-related Diabetes, ySIDD: Youth-onset Severe Insulin Deficient Diabetes, ySIRD: Youth-onset Severe Insulin Resistant Diabetes

**Supplementary Table 7. Descriptive characteristics of sex-specific clusters**

|  | **Male** |  |  | **Female** |  |  |
| --- | --- | --- | --- | --- | --- | --- |
|  | **yMOD** | **ySIDD** | **ySIRD** | **yMOD** | **ySIDD** | **ySIRD** |
| *N* | 122 (19.0%) | 48 (7.5%) | 98 (14.3%) | 189 (15.3%) | 72 (11.2%) | 112 (17.5%) |
| SEARCH | 34 (27.9%) | 41 (85.4%) | 59 (60.2%) | 52 (27.5%) | 53 (73.6%) | 65 (58%) |
| TODAY | 88 (72.1%) | 7 (14.6%) | 39 (39.8%) | 137 (72.5%) | 19 (26.4%) | 47 (42%) |
| Socio-demographics |  |  |  |  |  |  |
| Age at diagnosis |  |  |  |  |  |  |
| *<=13* | 25 (20.5%) | 21 (43.8%) | 28 (28.6%) | 95 (50.3%) | 27 (37.5%) | 49 (43.8%) |
| *14-15* | 37 (30.3%) | 8 (16.7%) | 23 (23.5%) | 35 (18.5%) | 20 (27.8%) | 23 (20.5%) |
| *>15* | 60 (49.2%) | 19 (39.6%) | 47 (48%) | 59 (31.2%) | 25 (34.7%) | 40 (35.7%) |
| Race-Ethnicity |  |  |  |  |  |  |
| *NH White* | 42 (34.4%) | 22 (45.8%) | 24 (24.5%) | 35 (18.5%) | 19 (26.4%) | 30 (26.8%) |
| *NH Black* | 25 (20.5%) | 12 (25%) | 33 (33.7%) | 55 (29.1%) | 30 (41.7%) | 50 (44.6%) |
| *Hispanic* | 47 (38.5%) | 10 (20.8%) | 30 (30.6%) | 80 (42.3%) | 17 (23.6%) | 24 (21.4%) |
| *NH Other* | 8 (6.6%) | 4 (8.3%) | 11 (11.2%) | 19 (10.1%) | 6 (8.3%) | 8 (7.1%) |
| BMI (kg/m^2^) | 33.2 (4.2) | 27.2 (7.2) | 38.4 (7.1) | 33.7 (3.4) | 30.6 (4.8) | 39.1 (7.4) |
| HbA1c (%) | 5.6 (5.3, 6.1) | 7.7 (6.6, 8.7) | 6.3 (5.9, 7.5) | 5.8 (5.4, 6.3) | 7.4 (5.8, 9.1) | 6.2 (5.8, 7.1) |
| Fasting glucose (mg/dL) ^a^ | 104.4 (19.6) | 119.1 (29.1) | 113.7 (25.6) | 100.4 (17) | 116.7 (25.4) | 115.8 (22.7) |
| Fasting Insulin (uU/mL) ^a^ | 25.1 (20.8) | 21.7 (11.3) | 43.7 (33.8) | 26.9 (16.7) | 19.4 (10.1) | 43.2 (22.1) |
| Fasting C-peptide (ng/mL) | 3.3 (1.3) | 1.7 (1.1) | 5 (2) | 3.7 (1.5) | 2.4 (1.1) | 5.1 (1.7) |
| LDL cholesterol (mg/dL) | 80.5 (23.2) | 110.6 (30.8) | 94 (26.2) | 80 (21.5) | 108.6 (29.9) | 97.1 (28.7) |
| HDL cholesterol (mg/dL) | 36.1 (7) | 48.4 (9.3) | 33.9 (7.9) | 37.8 (7.4) | 48.1 (9.8) | 40 (7.6) |
| Total cholesterol (mg/dL) | 139.2 (29.7) | 182.1 (39.2) | 160.7 (34.5) | 140.8 (26.7) | 181.4 (35.1) | 163.8 (38) |
| Triglycerides (mg/dL) ^a^ | 97.8 (59.5) | 68.1 (17) | 139.9 (98.9) | 103 (56.4) | 87.1 (45.9) | 127.5 (76.6) |
| Systolic BP (mmHg) | 112.1 (9.6) | 110.3 (10.1) | 125.4 (8.7) | 107.6 (8.1) | 109.2 (10.3) | 122.7 (9.8) |
| Diastolic BP (mmHg) | 64.6 (7.7) | 68.3 (9.2) | 75.7 (9.1) | 63.9 (7.2) | 67.5 (7.4) | 76.6 (9.2) |

^a^ Only available for TODAY (N = 337); yMOD: Youth-onset Mild Obesity-related Diabetes, ySIDD: Youth-onset Severe Insulin Deficient Diabetes, ySIRD: Youth-onset Severe Insulin Resistant Diabetes

**Supplementary Table 8. Missingness of MNSI variables by study, N = 501**

|  | Total | SEARCH | TODAY |
| --- | --- | --- | --- |
| Outcomes--MNSI: 9. Has your doctor ever told you that you have diabetic neuropathy? | 2 (0.4%) | 1 (0.5%) | 1 (0.3%) |
| Outcomes--MNSI: 11. Are your symptoms worse at night? | 1 (0.2%) | 0 | 1 (0.3%) |
| Outcomes--MNSI: 13. Are you able to sense your feet when you walk? | 1 (0.2%) | 1 (0.5%) | 0 |
| Outcomes--MNSI: 1a. Appearance of Feet, Right Normal? | 1 (0.2%) | 0 | 1 (0.3%) |
| Outcomes--MNSI: 1a. Appearance of Feet, Left Normal? | 1 (0.2%) | 0 | 1 (0.3%) |
| Outcomes--MNSI: 3. Ankle Reflexes (R) | 9 (1.8 %) | 5 (2.7%) | 4 (1.3%) |
| Outcomes--MNSI: 3. Ankle Reflexes (L) | 8 (1.6%) | 4 (2.2%) | 4 (1.3%) |
| Outcomes--MNSI: 4. Vibration perception at the great toe (R) | 4 (0.8%) | 2 (1.1%) | 2 (0.6%) |
| Outcomes--MNSI: 4. Vibration perception at the great toe (L) | 5 (1.0%) | 2 (1.1%) | 3 (0.9%) |
| Outcomes--MNSI: 5. 10 gm filament (number of applications detected out of 10 applications): (R) | 3 (0.6%) | 1 (0.5%) | 2 (0.6%) |
| Outcomes--MNSI: 5. 10 gm filament (number of applications detected out of 10 applications): (L) | 5 (1.0%) | 2 (1.1%) | 3 (0.9%) |

**Supplementary Table 9. Descriptive characteristics of the analytic sample stratified by identified subphenotypes (factorial diagnosis criteria)**

|  | **Total** | **yMOD** | **ySIDD** | **ySIRD** |
| --- | --- | --- | --- | --- |
| *N* | 722 | 338 (46.8%) | 160 (22.2%) | 224 (31.0%) |
| SEARCH | 385 (53.3%) | 108 (32%) | 146 (91.2%) | 131 (58.5%) |
| TODAY | 337 (46.7%) | 230 (68%) | 14 (8.8%) | 93 (41.5%) |
| Age at diagnosis (years) |  |  |  |  |
| *≤13* | 294 (40.7%) | 132 (39.1%) | 75 (46.9%) | 87 (38.8%) |
| *14-15* | 157 (21.7%) | 75 (22.2%) | 32 (20%) | 50 (22.3%) |
| *>15* | 271 (37.5%) | 131 (38.8%) | 53 (33.1%) | 87 (38.8%) |
| Female % | 722 (56.5%) | 338 (62.4%) | 160 (47.5%) | 224 (54%) |
| Race-Ethnicity |  |  |  |  |
| *NH White* | 226 (31.3%) | 88 (26%) | 81 (50.6%) | 57 (25.4%) |
| *NH Black* | 217 (30.1%) | 91 (26.9%) | 39 (24.4%) | 87 (38.8%) |
| *Hispanic* | 221 (30.6%) | 130 (38.5%) | 30 (18.8%) | 61 (27.2%) |
| *NH Other* | 58 (8%) | 29 (8.6%) | 10 (6.2%) | 19 (8.5%) |
| ***Key biomarkers for clustering*** |  |  |  |  |
| BMI (kg/m^2^) | 32.8 (7.8) | 33.3 (4.3) | 24.0 (6.1) | 38.5 (7.2) |
| HbA1c (%) | 6.1 (5.6, 7.1) | 5.8 (5.4, 6.3) | 7.3 (6.3, 8.5) | 6.2 (5.8, 7.3) |
| Fasting C-peptide (ng/mL) | 3.5 (2) | 3.5 (1.5) | 1.5 (1) | 5 (1.8) |
| LDL cholesterol (mg/dL) | 89.7 (27.2) | 82.1 (23.9) | 94.8 (27) | 97.5 (28.9) |
| HDL cholesterol (mg/dL) | 40.7 (10.4) | 37.8 (7.6) | 51.8 (10.8) | 37.1 (8.2) |
| Systolic BP (mmHg) | 113.3 (11.8) | 109.1 (9.2) | 108.3 (10.8) | 123.3 (9.7) |
| Diastolic BP (mmHg) | 68.6 (9.7) | 64.1 (7.3) | 68.2 (8.8) | 75.8 (9.2) |
| ***Other biomarkers*** |  |  |  |  |
| Fasting glucose (mg/dL) ^a^ | 106.5 (21.3) | 102.3 (18.3) | 124.8 (28.8) | 114 (23.7) |
| Fasting Insulin (uU/mL) ^a^ | 30.1 (22.2) | 25.4 (18) | 21.9 (10) | 43.1 (27.2) |
| Total cholesterol (mg/dL) | 154 (35) | 141.9 (30.1) | 165 (32.3) | 164.6 (37.9) |
| Triglycerides (mg/dL) ^a^ | 107.7 (67.1) | 99.9 (57.2) | 76.9 (34) | 131.6 (85.2) |
| ***DSPN*** |  |  |  |  |
| MNSI Measured | 546 (75.6%) | 285 (52.2%) | 96 (17.6%) | 165 (22.9%) |
| Abnormal Examination | 132 (24.2%) | 36 (27.3%) | 45 (34.1%) | 51 (38.6%) |
| Abnormal Questionnaire | 59 (10.8%) | 28 (47.5%) | 10 (16.9%) | 21 (35.6%) |
| Combined Abnormal | 165 (30.2%) | 56 (33.9%) | 47 (28.5%) | 62 (37.6%) |

^a^ Only available for TODAY (N = 337); yMOD: Youth-onset Mild Obesity-related Diabetes, ySIDD: Youth-onset Severe Insulin Deficient Diabetes, ySIRD: Youth-onset Severe Insulin Resistant Diabetes

Data are mean (SD) or median (IQR) for continuous variables and frequency (percentage) for categorical variables.

Factorial: Any of as DAA negative and insulin sensitive (≥8.15), DAA negative and insulin resistant (<8.15), Missing DAA and Insulin Resistant (<8.15), Missing DAA and Insulin Sensitive (≥8.15), or Missing Both DAA and insulin sensitivity.

**Supplementary Table 10. Descriptive characteristics of the analytic sample stratified by identified subphenotypes (initial provider diagnosis criteria)**

|  | **Total** | **yMOD** | **ySIDD** | **ySIRD** |
| --- | --- | --- | --- | --- |
| *N* | 640 | 299 (46.7%) | 140 (21.9%) | 201 (31.4%) |
| SEARCH | 303 (47.3%) | 89 (29.8%) | 95 (67.9%) | 119 (59.2%) |
| TODAY | 337 (52.7%) | 210 (70.2%) | 45 (32.1%) | 82 (40.8%) |
| Age at diagnosis (years) |  |  |  |  |
| *≤13* | 243 (38%) | 117 (39.1%) | 53 (37.9%) | 73 (36.3%) |
| *14-15* | 139 (21.7%) | 66 (22.1%) | 32 (22.9%) | 41 (20.4%) |
| *>15* | 258 (40.3%) | 116 (38.8%) | 55 (39.3%) | 87 (43.3%) |
| Female % | 640 (58.8%) | 299 (60.5%) | 140 (62.1%) | 201 (53.7%) |
| Race-Ethnicity |  |  |  |  |
| *NH White* | 158 (24.7%) | 79 (26.4%) | 28 (20%) | 51 (25.4%) |
| *NH Black* | 209 (32.7%) | 76 (25.4%) | 56 (40%) | 77 (38.3%) |
| *Hispanic* | 215 (33.6%) | 116 (38.8%) | 46 (32.9%) | 53 (26.4%) |
| *NH Other* | 58 (9.1%) | 28 (9.4%) | 10 (7.1%) | 20 (10%) |
| ***Key biomarkers for clustering*** |  |  |  |  |
| BMI (kg/m^2^) | 34.6 (6.5) | 33.3 (3.8) | 30.5 (5.7) | 39.2 (7.3) |
| HbA1c (%) | 6 (5.5, 6.8) | 5.7 (5.4, 6.2) | 6.5 (5.8, 8.1) | 6.3 (5.8, 7.3) |
| Fasting C-peptide (ng/mL) | 3.8 (1.8) | 3.4 (1.4) | 2.3 (1.1) | 5.3 (1.8) |
| LDL cholesterol (mg/dL) | 91.1 (28.2) | 79.2 (21.3) | 107.1 (30.5) | 97.6 (28.2) |
| HDL cholesterol (mg/dL) | 39.2 (9.2) | 36.8 (6.9) | 48.1 (9.4) | 36.6 (8.1) |
| Systolic BP (mmHg) | 114.3 (11.6) | 109 (9.3) | 112.1 (9.9) | 123.5 (10.1) |
| Diastolic BP (mmHg) | 69 (9.8) | 64.2 (7.5) | 70 (8.4) | 75.7 (9.5) |
| ***Other biomarkers*** |  |  |  |  |
| Fasting glucose (mg/dL) ^a^ | 106.5 (21.3) | 101.6 (18.2) | 114 (25.7) | 114.8 (22.6) |
| Fasting Insulin (uU/mL) ^a^ | 30.1 (22.2) | 25.8 (18.3) | 21.5 (11.1) | 45.8 (27.6) |
| Total cholesterol (mg/dL) | 155.1 (36.2) | 138.4 (26.6) | 177.1 (36.3) | 165.3 (37.1) |
| Triglycerides (mg/dL) ^a^ | 107.7 (67.1) | 99.5 (57.8) | 86.9 (42.9) | 140.1 (87) |
| ***DSPN*** |  |  |  |  |
| MNSI Measured | 489 (76.4%) | 252 (51.5%) | 91 (18.6%) | 146 (29.9%) |
| Abnormal Examination | 102 (20.9%) | 28 (27.5%) | 25 (24.5%) | 49 (48.0%) |
| Abnormal Questionnaire | 54 (11.0%) | 24 (44.4%) | 10 (18.5%) | 20 (37.0%) |
| Combined Abnormal | 134 (27.4%) | 46 (34.3%) | 28 (20.9%) | 60 (44.8%) |

^a^ Only available for TODAY (N = 337); yMOD: Youth-onset Mild Obesity-related Diabetes, ySIDD: Youth-onset Severe Insulin Deficient Diabetes, ySIRD: Youth-onset Severe Insulin Resistant Diabetes

Data are mean (SD) or median (IQR) for continuous variables and frequency (percentage) for categorical variables.

Provider: The initial provider classification of all T2DM and unknown diabetes as T2DM. Second, we identified T2DM as, based on etiologic evidence.

**Supplementary Table 11. Cross-sectional association of subphenotype with distal symmetrical polyneuropathy**

|  | Subphenotype | Examination | Questionnaire | Combined |
| --- | --- | --- | --- | --- |
| Main clusters | yMOD (reference) | Ref (1.00) | Ref (1.00) | Ref (1.00) |
|  | ySIDD | 2.58 (1.74, 3.81) | 0.76 (0.37, 1.58) | 1.89 (1.35, 2.63) |
|  | ySIRD | 2.02 (1.4, 2.93) | 1.01 (0.59, 1.73) | 1.72 (1.28, 2.31) |
| Complete cases | yMOD (reference) | Ref (1.00) | Ref (1.00) | Ref (1.00) |
|  | ySIDD | 3.11 (2.02, 4.8) | 0.77 (0.37, 1.59) | 1.99 (1.39, 2.85) |
|  | ySIRD | 2.27 (1.51, 3.42) | 0.96 (0.55, 1.66) | 1.74 (1.26, 2.39) |
| SEARCH-only sample | yMOD (reference) | Ref (1.00) | Ref (1.00) | Ref (1.00) |
|  | ySIDD | 1.11 (0.81, 1.51) | 0.62 (0.27, 1.42) | 1.11 (0.83, 1.49) |
|  | ySIRD | 1.06 (0.79, 1.44) | 0.75 (0.37, 1.51) | 1.11 (0.85, 1.46) |
| Auto-antibody based definition for SEARCH + TODAY | yMOD (reference) | Ref (1.00) | Ref (1.00) | Ref (1.00) |
|  | ySIDD | 2.65 (1.86, 3.78) | 0.9 (0.45, 1.77) | 2.04 (1.51, 2.75) |
|  | ySIRD | 1.88 (1.33, 2.67) | 1.18 (0.69, 2) | 1.66 (1.25, 2.21) |
| Initial provider classification for SEARCH + TODAY | yMOD (reference) | Ref (1.00) | Ref (1.00) | Ref (1.00) |
|  | ySIDD | 1.98 (1.27, 3.09) | 1.05 (0.53, 2.09) | 1.55 (1.06, 2.25) |
|  | ySIRD | 2.22 (1.51, 3.26) | 1.25 (0.71, 2.19) | 1.9 (1.4, 2.57) |

Etiologic: Classification based on etiologic evidence by expert adjudicators

Factorial: Any of as DAA negative and insulin sensitive (≥8.15), DAA negative and insulin resistant (<8.15), Missing DAA and Insulin Resistant (<8.15), Missing DAA and Insulin Sensitive (≥8.15), or Missing Both DAA and insulin sensitivity.

Provider: The initial provider classification of all T2DM and unknown diabetes as T2DM. Second, we identified T2DM as, based on etiologic evidence.

Associations are prevalence ratios from marginal structural Poisson regressions using multiple imputation for missing covariates and inverse probability weights for non-participation in Michigan Neuropathy Screening Instrument assessment. All associations in main clusters, complete cases analysis and SEARCH-only are adjusted for age category, sex and race-ethnicity.

**Supplementary Table 12. Adult-onset T2DM Study Comparison**

|  | GDS | | | ANDIS | | |
| --- | --- | --- | --- | --- | --- | --- |
|  | MOD | SIDD | SIRD | MOD | SIDD | SIRD |
| BMI (kg/m^2^) | 34.7 (6.4) | 27.0 (3.7) | 34.2 (4.5) | 35.7 (5.4) | 28.8 (4.8) | 33.8 (5.2) |
| HbA1c (%) | 6.5 (0.9) | 8.7 (1.3) | 6.2 (0.7) | 7.4 (3.6) | 11.8 (3.9) | 7.1 (3.6) |

Abbreviations: GDS, German Diabetes Study; ANDIS, All New Diabetics in Scania

**Supplementary Figure 1. Flowchart of analytic sample**

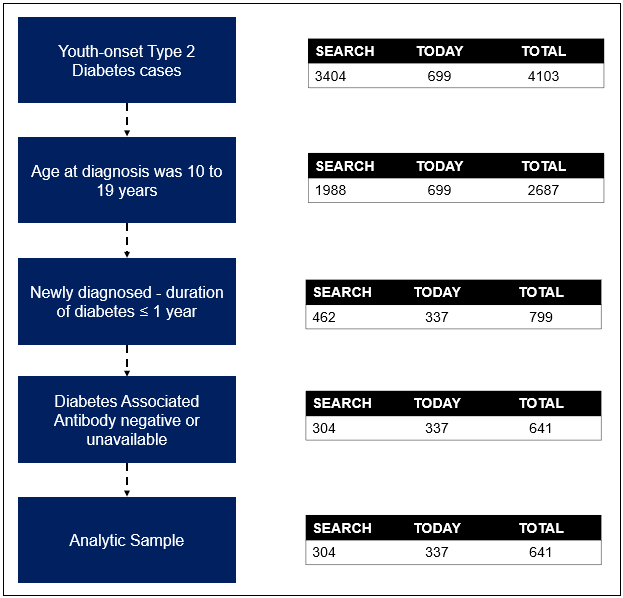

Abbreviation: GAD, Glutamic Acid Decarboxylase

**Supplementary Figure 2. Elbow plot for K-means clustering**

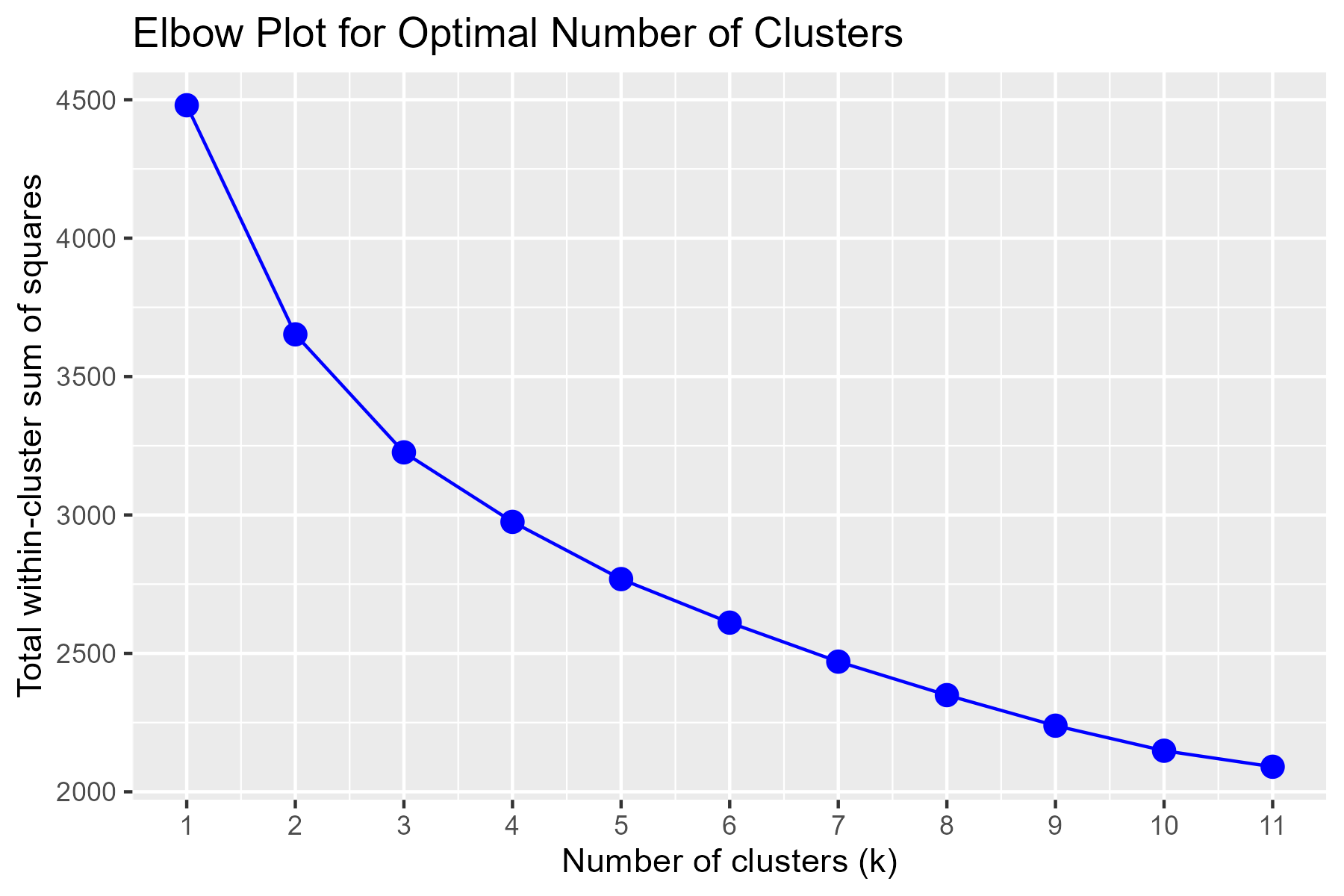

**Supplementary Figure 4. Mean and SD of variables from clustering within subset and diagnosis criteria**

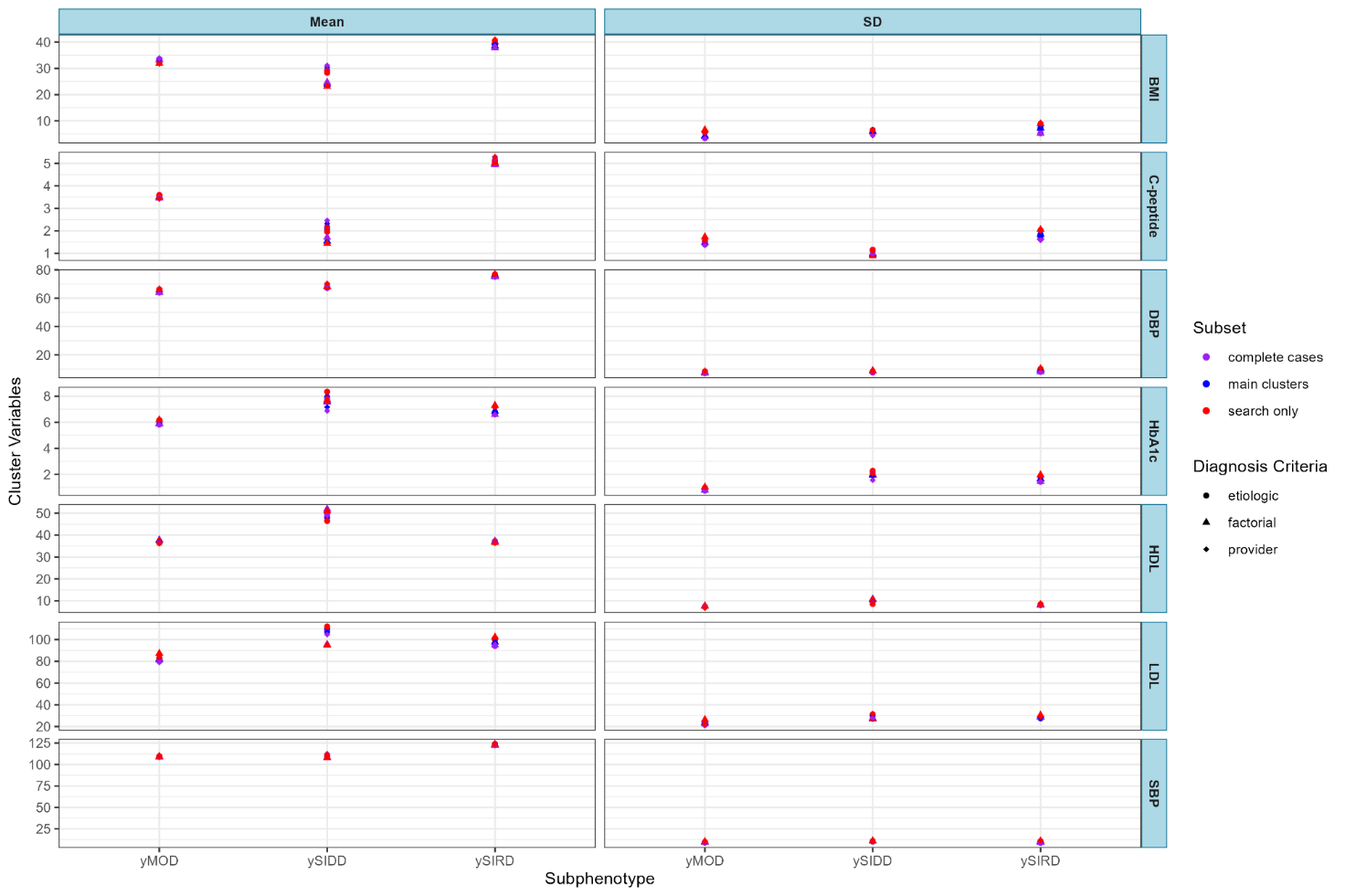

Etiologic: Classification based on etiologic evidence by expert adjudicators

Factorial: Any of as DAA negative and insulin sensitive (≥8.15), DAA negative and insulin resistant (<8.15), Missing DAA and Insulin Resistant (<8.15), Missing DAA and Insulin Sensitive (≥8.15), or Missing Both DAA and insulin sensitivity.

Provider: The initial provider classification of all T2DM and unknown diabetes as T2DM. Second, we identified T2DM as, based on etiologic evidence.

**Supplementary Figure 5. Prevalence Ratio of abnormal neuropathy symptoms by subset**

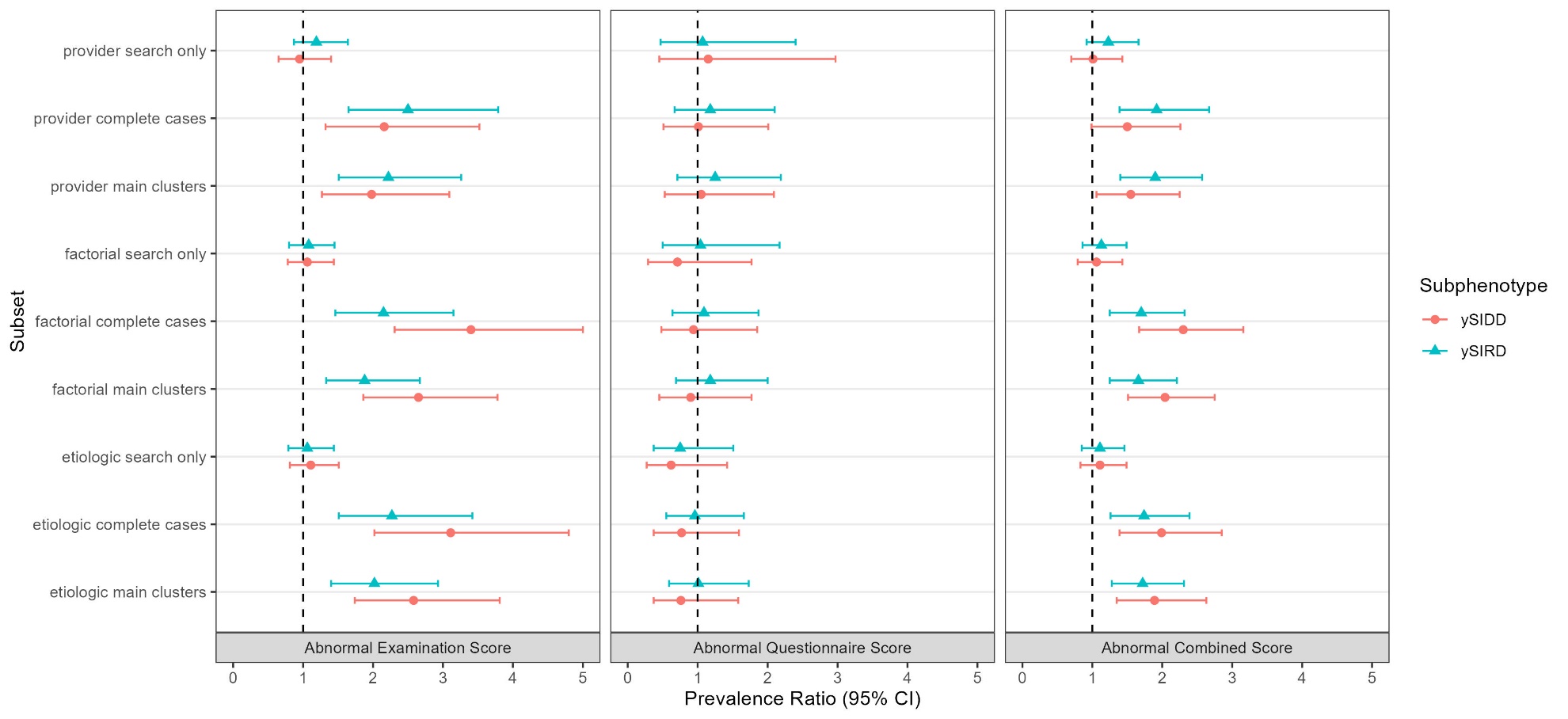

Etiologic: Classification based on etiologic evidence by expert adjudicators

Factorial: Any of as DAA negative and insulin sensitive (≥8.15), DAA negative and insulin resistant (<8.15), Missing DAA and Insulin Resistant (<8.15), Missing DAA and Insulin Sensitive (≥8.15), or Missing Both DAA and insulin sensitivity.

Provider: The initial provider classification of all T2DM and unknown diabetes as T2DM. Second, we identified T2DM as, based on etiologic evidence.
